# The social determinants associated with decreased rate of breast cancer screening, delayed presentation, and advanced stage diagnosis of breast cancer: A systematic review

**DOI:** 10.1101/2023.09.22.23295953

**Authors:** Madonna A. Fekry, Justin C. Yang

## Abstract

**Background:** There are many barriers that hinder breast cancer (BC) early detection such as social, demographic, and economic factors. We examined the barriers to early detection.

**Methods:** PubMed, Scopus, and Web of Science databases were chosen to conduct a comprehensive literature search. The Preferred Reporting Items for Systematic reviews and Meta-Analyses (PRISMA) was used to select the relevant studies on decreased rate of BC screening, delayed presentation of BC, and advanced stage diagnosis of BC.

**Results:** The literature demonstrates that several determinants had a significant impact on the delay in seeking medical help, rate of performing breast cancer screening (BCS), and stage at diagnosis of BC. Younger age, rural residence, being non-white, being single, low socioeconomic status, absence of medical insurance, having no paid job, low educational level, positive family history of BC, and having TNBC or HER2E BC subtypes were significantly associated with presenting at advanced stages, decreased rate of BCS, and delayed presentation. Meanwhile, the associations between BC and BMI, parity, religion, and menopausal status were underexamined in the literature.

**Conclusion:** Promoting early detection of BC should be taking the sociodemographic disparities into consideration. To address these disparities, raising public awareness, implementing universal health coverage (UHC), and increasing government expenditure on health and education are needed, especially among vulnerable societies.

## Introduction

Breast cancer (BC) is the most common cancer and the second leading cause of cancer death among women worldwide. In 2020, there were 2.3 million new breast cancer cases and 685,500 breast cancer related deaths globally (1). Unlike some other cancers, there are no known bacterial or viral infections linked to its development. BC has many risk factors. However, approximately half of BC patients have no identifiable BC risk factors. There are certain factors that are associated with increased risk of developing BC such as being a woman, having certain mutations such as BRCA1/2, increasing age, family history of BC, being overweight or obese, and using hormonal replacement therapy (HRT). Disappointingly, controlling all these risk factors is likely to only reduce the risk of developing BC by only 30% (1). Consequently, early detection of BC is crucial; early-stage BC is curable, while advanced stage BC is associated with poor prognosis. However, there are many barriers that hinder BC early detection such as social, demographic, and economic factors. These factors can limit patients’ access to effective, affordable, and timely breast cancer services (2). Therefore, the aim of this systematic review is to examine the determinants associated with decreased rate of BC screening, delayed presentation, and advanced stage diagnosis of BC.

## Methods

### Search strategy

The Preferred Reporting Items for Systematic reviews and Meta-Analyses (PRISMA) guidelines were followed to select the relevant studies on decreased rate of breast cancer screening, delayed presentation, and advanced stage diagnosis of breast cancer. The inclusion criteria were full-text studies, quantitative studies, and studies linked to either BC staging or BC screening. The exclusion criteria were studies older than 10 years, qualitative studies, and studies linked to BC awareness. PubMed, Scopus, and Web of Science databases were chosen to conduct a comprehensive literature search. The search strategy was established using a combination of keywords and Medical Subject Headings (MeSH) terms. Appendix 1 displays the keywords that have been used in the research strategy. Boolean operators (AND & OR) were used to combine keywords in the literature search. Breast neoplasms/ diagnosis and social determinants of health MeSH terms were applied to the advanced search of PubMed database.

### Data extraction

The data extraction was carried out using a data extraction Excel sheet. The extracted data were sample size, country, study design, study outcomes (e.g., advanced or late-stage diagnosis of BC, BC screening, and delayed presentation), and exposure variables (e.g., age, residence (rural vs. urban), race, BMI, marital status, religion, education status, socioeconomic status, employment status, family history, parity, BC subtype, and menopausal status).

### Quality assessment of the eligible articles

The quality of the eligible articles was assessed by MAF and JCY. A quality assessment tool for observational research was used to assess the potential bias in each included study. The tool included nine questions measuring the appropriateness and clarity of the study question, objectives, study variables, statistical analysis, dealing with confounders, and sample size. For each positive answer on each question, a score of one was given and for each negative or not available answer, a score of zero was given. The overall quality score of an article was expressed as the sum of scores of each study. Zero represents the lowest quality score and nine represents the highest quality score. The higher the score, the higher quality of the article and the lower the score, the lower quality of the article. In this systematic review, sixteen studies were given a quality score of nine, twenty-four studies were given a quality score of eight, thirteen studies were given a quality score of seven, and five studies were given a quality score of six (Appendix 2).

### Search results

Figure 1 displays the PRISMA flow chart for the search and screening strategy. The searches were conducted from December 2022 to January 2023. Using the advanced search builder of PubMed, Scopus, and Web of Science, a search for the keywords (Appendix 1) was conducted, resulting in 667, 7, and 6 related articles, respectively. The total number of articles identified in the review was 680. After applying MeSH terms, 427 articles were excluded before screening. Moreover, twenty-three duplicate articles were removed before screening. Out of the 230 articles that have been screened, 150 articles were excluded for being not linked to either BC staging or BC screening. Sixteen qualitative studies and six studies related to BC awareness were excluded, resulting in fifty-eight studies included in this systematic review.

**Figure 1:**
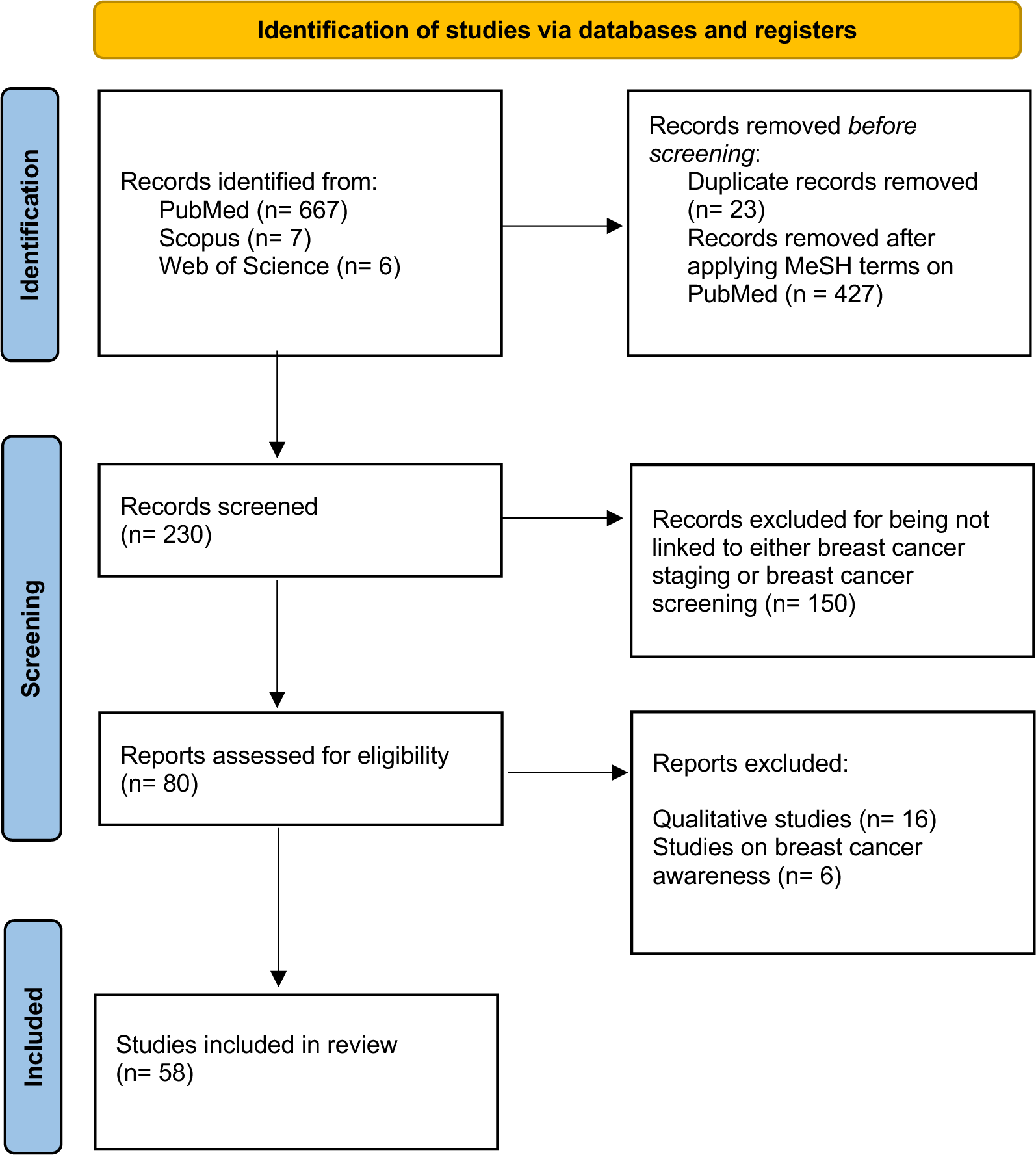
PRISMA flow chart of literature search and screening.

### Study characteristics

The study design of the fifty-eight papers was observational. There were 11 studies in the USA, five in Brazil, four in India, three in Nigeria, two in eight different countries in Sub-Saharan Africa, two in Uganda, two in Pakistan, two in Ethiopia, two in New Zealand, two in Asia, two in Thailand, one in 14 low-resource countries, one in Africa, one in Australia, one in Colombia, one in France, one in Gaza, one in Germany, one in Indonesia, one in Iran, one in Korea, one in Lesotho, one in Malaysia, one in Malta, one in Vietnam, one in Canada, one in Rwanda, one in Norway, one in Tunisia, one in south Africa, one in Spain, and one in Surabaya. The median sample size for the fifty-eight included studies was 1,643 participants (range: 120 – 1,400,000) (Appendix 2).

### Findings

The literature demonstrates that several determinants had a significant impact on the delay in seeking medical help, rate of performing breast cancer screening (BCS), and stage at diagnosis of BC. Extensive evidence from the literature demonstrated that there was a significant association between early stage of BC diagnosis and the increased rate of practicing BCS techniques such as breast self-examination (BSE), clinical breast examination (CBE), and mammography. Five studies with different study designs (one in Nigeria, one in North-East Ethiopia, one in Africa, one in Asia and one in Iran) have corroborated this claim (3–7). Four studies (one in Pakistan, one in Tunisia, one in Rwanda, and one in Uganda) observed that longer diagnostic journey was significantly associated with late-stage diagnosis of BC (8–11).

### Rate of performing BC screening

There was extensive evidence from the literature that age is highly associated with the rate of performing BCS techniques. Nine studies (two in the USA, one in Australia, one in Sub-Saharan Africa, one in Lesotho, one in Gaza, one in Thailand, one in Surabaya, and one in India) concluded that older individuals were significantly more likely to practice BCS techniques compared to younger individuals(12–20). These nine studies had different study designs and a wide range of number of participants (from 370 to 336,777). On the other hand, there was one cross sectional survey that has been conducted in India that observed a controversial association between age and rate of BCS performance(21). Increasing age was significantly associated with higher odds of performing BSE in Fiji but not significantly associated with performing BSE in Kashmir. The nonsignificant association observed in Kashmir could be due to the political strife in the region resulting in decreased access of all age groups to healthcare.

Five studies (one in the USA, one in fourteen low resources countries, one in Thailand, one in Germany, and one in Uganda) assessed the impact of urbanicity on rate of performing BCS techniques. Individuals who live in rural areas were less likely to practice BC screening compared to those who live in urban areas (12,17,22–24). Different study designs were used among these five studies such as cross-sectional studies, cohort studies, and surveys with a wide range of sample sizes (from 401 to 140,974). This confirms that the association between urbanicity and rate of BCS performance is significant regardless of the number of participants, the country, and the study design used.

Race, body mass index (BMI), marital status, and monthly income had a significant impact on the rate of BCS performance. Only one cross-sectional study (in three US Midwest states) assessed the association between race and rate of BCS performance. Non-white individuals were approximately half as likely to complete cancer screening compared to white individuals (12). One cross-sectional study (in 14 low resource countries) concluded that obese individuals had significant lower odds of utilizing BCS services compared to normal weight individuals (22). This study had good generalizability and large sample size (140,974). Another secondary data analysis (in India) concluded that obese women were more likely to undergo breast screening (20). However, secondary analyses may have some sort of bias since the investigators did not have complete control over the quality of data. Majority of studies (two in India, one in Australia, and one in Thailand) observed that married women were more likely to practice BCS compared to single women (13,17,20,21). However, another systematic review that included 70 studies concluded that marital status was not significantly associated with participation in mammography screening in Asia (25). Despite the high reliability of systematic reviews, the included studies in this systematic review had limited sample sizes which reduces its reliability and generalizability. Two studies in high income countries (Germany and Canada) and other two studies in low-middle income countries (Sub-Saharan Africa and 14 low resource countries) concluded that individuals with high income and socioeconomic status had significant higher rates of utilizing BC services compared to those with low income and socioeconomic status(14,22,23,26). Only one questionnaire-based study (in Northern Vietnam) that concluded that high income was associated with low rate of utilizing BC services (27). Besides the fact that questionnaires could have had response bias, this study had limited sample size (306) which reduces its reliability.

A majority of studies that have been conducted in many different countries and regions revealed the association between poor utilization of BCS techniques and education inequalities. Highly educated individuals were significantly more likely to undergo BCS compared to individuals with no or low level of education (14,15,17–20,22,28,29). In contrast, one questionnaire-based study (in Germany) concluded that individuals with middle or high education were significantly less likely to undergo mammography screening compared to individuals with low educational status (23). Because of being a questionnaire-based study, this study implied a risk of response distortions and incorrect answers.

The association between employment status and rate of BCS performance was controversial in the literature. Two studies (one in Germany and one in Asia) concluded that there was no significant association between rate of BCS performance and employment status (23,25). However, these two studies lack reliability because of the risk of response bias of one of them and limited sample size of the other. One cross sectional study with good sample size (in the USA) concluded that unemployment was significantly associated with low rate of screening (18). In contrast, another secondary analysis (in Australia) concluded that unemployed women had higher odds of practicing annual CBE (13). Despite the good sample size used in this secondary analysis, the researchers claimed that reliance on self-reported measures could have confounded their results.

Family history and having health insurance coverage were significantly associated with the rate of BCS performance in the literature. Three cross-sectional studies (one in Gaza, one in Ethiopia, and one in Surabaya) concluded that women with positive family history of BC had significant higher odds of utilizing BCS services compared to women with no family history (16,19,28). Four studies (one in four Sub-Saharan Africa, one in 14 low resource countries, one in Thailand, and one in the USA) concluded that women with health insurance coverage were significantly more likely to undergo BCS compared to those with no health insurance (14,17,18,22). These four studies included large sample sizes (from 1541 to 140,974) which gives them high level of reliability. On the other hand, two other studies with relatively smaller sample sizes (one in Lesotho and one in Germany) concluded that health insurance coverage was not significantly associated with performing BC screening (15,23).

### Delay in seeking medical help, diagnosis, or presentation

The association between age and delay in seeking medical help was controversial in the literature. Most studies that have been conducted in Pakistan, Rwanda, Nigeria, and Uganda observed no significant association between age and delay in presentation or diagnosis (8,9,11,30). The nonsignificant association could not be observed because of the limited sample size of these studies. Nevertheless, two other cohort studies that had relatively larger sample sizes (one in Sub-Saharan Africa and one in Iran) concluded that older patients were significantly more likely to have delayed diagnosis (7,31).

Three studies (one in Tunisia, one in Iran, and one in India) concluded that rural residents had longer delay in seeking medical advice compared to urban residents (7,9,10,32).

Marital status and monthly income were associated with the delayed presentation or diagnosis. Two studies (one cohort in five Sub-Saharan African countries and one cross-sectional in Nigeria) concluded that unmarried patients were more likely to have delayed presentation or diagnosis compared to married patients (30,31). In contrast, one cohort study (in Pakistan) concluded that unmarried patients had shorter delay in seeking medical help compared to married patients (9). This could be explained as married women in Pakistan may fear getting divorced after getting diagnosed with BC. Five different study designs (two in Pakistan, one in Iran, one in five Sub-Saharan African countries, and one in India) concluded that low income and socioeconomic status was significantly associated with longer delay in diagnosis compared to high income and socioeconomic status (7,9,31–33).

Low level of education was significantly associated with longer delay in presentation. Several researchers have corroborated this claim (3,8,31–33). However, two other studies (one in Pakistan and one in Nigeria) claimed that there was no significant association between level of education and length of diagnostic journey (9,30). These two studies did not perform any multilevel analyses to adjust for possible confounders.

There was not enough evidence in the literature on the impact of employment status and parity on the length of diagnostic journey. One cohort study (in Pakistan) concluded that working patients had shorter delay than nonworking patients (9). Two other cross-sectional studies (in Nigeria and Uganda) concluded that employment status was not significantly associated with delayed presentation (11,30). However, in the three studies, researchers did not adjust for possible confounders such as monthly income. Only one case control study (in Tunisia) concluded that having more than three dependents was significantly associated with longer delay in seeking consultation (10).

One case-control study (in Pakistan) concluded that individuals with positive family history of BC were significantly less likely to delay their first consultation compared to those with negative family history (33). In contrast, another questionnaire-based study (in Indonesia) concluded that having family history of cancer was associated with increased odds of delay in diagnosis in Indonesia (34). However, this study had lack of reliability because of its limited sample size and possible response bias of participants. Nonetheless, three other studies (one questionnaire based in Rwanda, one cross-sectional in Brazil, and one cross-sectional in Uganda) claimed that there was no significant association between family history and delay in seeking consultation (8,11,35).

### Advanced or late-stage diagnosis of BC

Extensive evidence from the literature demonstrated that younger patients were more likely to get diagnosed at advanced stages compared to older patients. Eight studies with different study designs that have been conducted in different countries and regions, have corroborated this claim. These studies had good generalizability with large sample sizes that ranged from 2,888 to 195,201. In contrast, other three studies with much less sample sizes (one in Thailand, one in Colombia and one in Korea) concluded that older patients had higher odds of getting diagnosed at late stages compared to younger patients (36–38). Moreover, four other studies that have been conducted in three African countries (two in Nigeria, one in South Africa, and one in Ethiopia) concluded that age was not significantly associated with advanced stage diagnosis of BC (4,5,39,40).

Four studies (one in Iran, one in New Zeeland, one in Nigeria, and one in North-west Ethiopia) concluded that rural residents had higher odds of presenting with late-stage BC compared to urban residents (4,7,40,41). However, three other studies (one cohort in the USA, one cohort in Ohio, and one population-based in Korea) observed no significant association between urbanicity and advanced or late-stage diagnosis of BC (38,42,43). This nonsignificant association could be explained that rural areas in the USA and Korea are not deprived compared to rural areas in other countries.

The literature demonstrated extensive evidence on the association between race and advanced stage diagnosis of BC in the USA. Six studies concluded that black patients had higher odds to present with advanced stage diagnosis compared to white patients (43–48).

Two studies with large sample sizes (one in Brazil and one in Ohio) claimed that unmarried patients were more likely to get diagnosed at advanced stages compared to married patients (43,49). However, other three survey-based studies (one in Nigeria, one in South Africa and one North-west Ethiopia) observed no significant association between marital status and advanced stage diagnosis of BC (4,5,39). Besides the fact that these survey-based studies could have had response bias, they had small sample sizes.

There was an uncertainty in the literature regarding the impact of religion, employment status, and family history of BC on the advanced stage diagnosis of BC. One study (in Nigeria) concluded that Muslims had significant lower odds of presenting with advanced stages compared to Christians in Nigeria (40). However, the authors claimed that this finding could be explained as Muslims in their studies may belong to higher socioeconomic class. Conversely, another study (in North-west Ethiopia) concluded that Muslims were more likely to present with advanced disease compared to Christians (4). Only two survey-based studies (one in North-west Ethiopia and one in south Africa) that measured the association between employment status and stage at diagnosis, but they claimed no significant association between both variables (4,39). One study (in Iran) concluded that patients with positive family history of BC were more likely to get diagnosed at advanced stages (7). Having an aggressive disease could justify this conclusion as patients with family history could have certain mutations such as BRCA1/2 that are linked to aggressive disease. Another study (in North-west Ethiopia) observed no significant association between both variables (39).

One cross-sectional study (in Thailand) concluded that low family income was significantly associated with late-stage diagnosis of BC (36). However, two other studies with relatively larger sample sizes (in Mississippi and in New Zealand) observed no significant difference in advanced stage percentages among different deprived groups (48,50).

Five studies (two in Brazil, one in Thailand, and one in South India, and one in Nigeria) concluded that patients with no or low level of education were more likely to get diagnosed at advanced stages compared to patients with high of education (36,40,44,49,51). Conversely, two other survey-based studies (one in Nigeria and one in South Africa) observed no significant association between level of education and stage at diagnosis of BC (5,39). Besides the fact that these two studies could have had response bias, they did not adjust for any possible confounders using multilevel analyses.

Two studies with large sample sizes (one cross-sectional in the USA and one cohort in Virginia) concluded that patients with no medical insurance were more likely to present with advanced disease compared to those with medical insurance (45,47). In contrast, another study (in Mississippi) concluded that patients with medical insurance were more likely to present with late-stage BC (48). However, race could be a major confounder in this study.

One meta-analysis (in Africa) observed that triple negative or HER2 enriched BC subtypes were more likely to present with advanced disease compared to other subtypes (3). Another prospective study (in New Zealand) observed that patients with ER and PR negative and HER2 positive were more likely to present with advanced or metastatic disease compared to patients with ER and PR positive and HER2 negative (41). Only one cross-sectional study with limited sample size (in Thailand) observed that postmenopausal status was significantly associated with late-stage diagnosis of BC (36).

## Discussion

This systematic review highlights the socioeconomic inequalities associated with decreased rate of breast cancer screening, delayed presentation, and advanced stage diagnosis of breast cancer. Younger age, rural residence, being non-white, being single, low socioeconomic status, absence of medical insurance, having no paid job, low educational level, positive family history of BC, and TNBC or HER2E BC subtypes were significantly associated with presenting at advanced stages, decreased rate of BCS, and delayed presentation. Furthermore, BMI, parity, religion, and menopausal status were underexamined in the literature.

Most of included studies elucidated that older individuals were significantly more likely to practice BCS and present with early disease compared to younger individuals (3,12–19,41–46,52). Women think that BC could only be developed at old age which explains why rate of practicing BCS is higher among older women compared to younger ones. Therefore, it is essential to assess the knowledge of females on BC and its risk factors and correct any misconceptions that may contribute to delayed diagnosis.

Most included studies observed a significant association between urbanicity and BC stage at diagnosis, rate of practicing BCS, and length of delay in seeking medical help. Individuals who reside in rural areas were more likely to present at advanced stages and have longer delay in seeking medical help and less likely to practice BCS compared to individuals who reside in urban areas (4,7,9,10,12,17,22–24,32,40,41). This association can be justified by the lack of awareness and education of people living in rural areas. Moreover, rural residents are more likely to use traditional methods of treatment due to long distances to specialized medical facilities. Therefore, increasing expenditure on education and health would help decrease the inequalities between rural and urban residents.

Race had a significant impact on rate of practicing BCS and stage of BC diagnosis in majority of included studies. Non-white individuals were less likely to perform BCS and present at early stages compared to white individuals (43–48). This can be explained by the discrimination from which the non-white individuals are suffering to access better education, jobs, and healthcare services. Therefore, it is mandatory to work on better strategies to eradicate racial disparities in schools, companies, and healthcare providers.

Most of the included studies elucidated significant differences in BC stage at diagnosis, BCS rate, and diagnostic journey between married and unmarried patients. Married patients are less likely to get diagnosed at advanced stages, have delayed presentation, and not practice BCS compared to unmarried patients (13,17,20,21,30,31,43,49). This can be explained by the fact that supportive spouses play a key role in encouraging the patients to seek medical help whenever needed. These findings demonstrate the importance of the social and psychological support that unmarried patients would need from the oncologists and other care providers (53). In contrast, there was only one study in Pakistan which claimed that unmarried patients had shorter delay in seeking medical help compared to married patients (9). This can be explained through Pakistani women fearing separation from their spouses.

Individuals with high income and socioeconomic status had significant higher rates of utilizing BC services and shorter delay in diagnosis compared to those with low income and socioeconomic status in most of included studies (7,9,14,22,23,26,31–33). This can be explained through the fact that individuals with low income and socioeconomic status tend to have less comprehensive medical insurance coverage compared to those with high income and socioeconomic status. Implementation of universal healthcare coverage (UHC) would help reduce barriers of care among individuals with low socioeconomic status. Additionally, providing complementary services that are free or of low cost would help improve access to care (54). These strategies would also help reduce the barriers to healthcare access caused by not having medical insurance. Two studies examined that patients with medical insurance were less likely to present at advanced stages compared to those with no medical insurance (45,47).

It was expected that patients with no paid job may not prioritize utilizing BCS services or access to medical facilities due to low income. However, the association between BC stage at diagnosis and employment status was underexamined in the literature. There were only two studies that have been conducted in Africa and Ethiopia observed no significant association between employment status and stage at diagnosis (4,39). Additionally, the association between employment status and practicing BCS was controversial; two studies observed no significant association, one study concluded that unemployment was significantly associated with low rate of BCS, and one another study concluded that unemployed women were more likely to practice BCS (13,18,23,25). The association between employment status and length of delay in seeking medical help was also controversial; two studies observed no significant association and one study concluded that working patients had shorter delay than nonworking patients (9,11,30).

Education inequalities were significantly associated with rate of BCS performance, diagnostic journey, and stage of BC diagnosis. Several studies claimed that highly educated individuals were more likely to practice BCS, have shorter delay, and were less likely to present at advanced stages compared to those with low educational level (5,8,14,15,17–20,22,28,29,31–33,36,40,44,49,51). This could be explained by the fact that individuals with low educational level are less likely to be offered good paying jobs; subsequently, they cannot afford access to healthcare. Therefore, increasing the government expenditure on education systems is the best strategy to reduce the education disparities among vulnerable individuals.

Majority of included studies that examined the association between family history and rate of BCS performance concluded that women with positive family history of BC were more likely to utilize BCS services compared to women with no family history (16,19,28). This can be explained by the fact that patients who have relatives with breast cancer are more attentive to its risk factors, symptoms, and BCS techniques. However, the association between family history and BC stage at diagnosis was controversial in the literature. One study in Iran concluded that positive family history was associated with advanced disease (7). Moreover, another study in Ethiopia observed no significant association between stage at diagnosis and family history of BC (39).

Patients with TNBC or HER2E subtypes were more likely to present at advanced stages compared to other subtypes. This finding was concluded by two studies which have been conducted in Africa and New Zealand (3,41). The reason behind this finding is that these subtypes have worse prognosis and are more aggressive than luminal A, B1, and B2 subtypes which explains why patients present with advanced disease.

The findings of this systematic review revealed the social, racial, economic, and demographic disparities associated with decreased rate of BC screening, delayed presentation, and advanced stage diagnosis of BC in several countries. To address these disparities, raising public awareness, implementing UHC, and increasing government expenditure on health and education are needed. This systematic review has some limitations. It could have introduced selection bias while selecting the relevant articles. Moreover, the assessment of study quality was performed using a general tool for observational studies not a specific tool for each study design. It is recommended to examine the association between advanced stage diagnosis of BC and sociodemographic variables, such as gender, religion, and parity in future research.

## Conclusion

This systematic review highlights the socioeconomic inequalities associated with decreased rate of breast cancer screening, delayed presentation, and advanced stage diagnosis of breast cancer. Younger age, rural residence, being non-white, being single, low socioeconomic status, absence of medical insurance, having no paid job, low educational level, positive family history of BC, and TNBC or HER2E BC subtypes were significantly associated with presenting at advanced stages, decreased rate of BCS, and delayed presentation. To address these disparities, raising public awareness, implementing UHC, and increasing government expenditure on health and education are needed.

## Data Availability

All data produced in the present study are available upon reasonable request to the authors

## Abbreviations

BC: breast cancer
HER2E: HER2 enriched
TNBC: triple-negative breast cancer
BMI: body mass index
BCS: breast cancer screening
UHC: universal healthcare coverage

## Authors’ contributions

MAF and JCY contributed to developing study concept and design.

MAF and JCY contributed to searching the databases and assessing the quality of included studies.

MAF and JCY contributed to writing the initial draft of the manuscript.

JCY contributed to reviewing the whole systematic review.

MAF and JCY approved the final version of the manuscript.

## Appendices

**Appendix 1.**
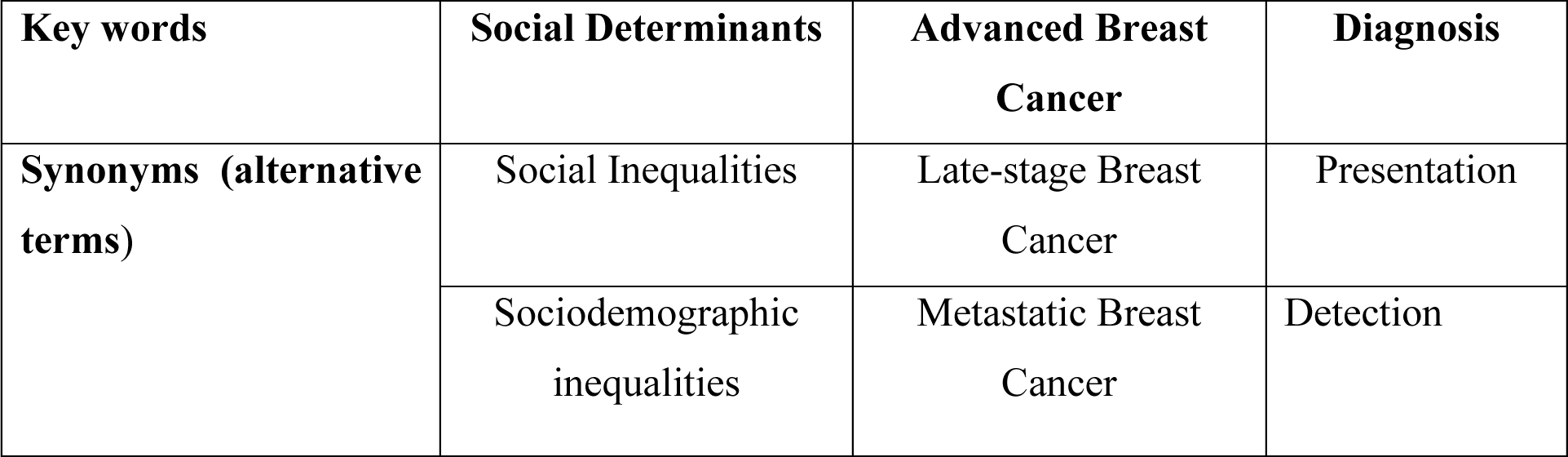
Keywords used in the literature search.

**Appendix 2.**
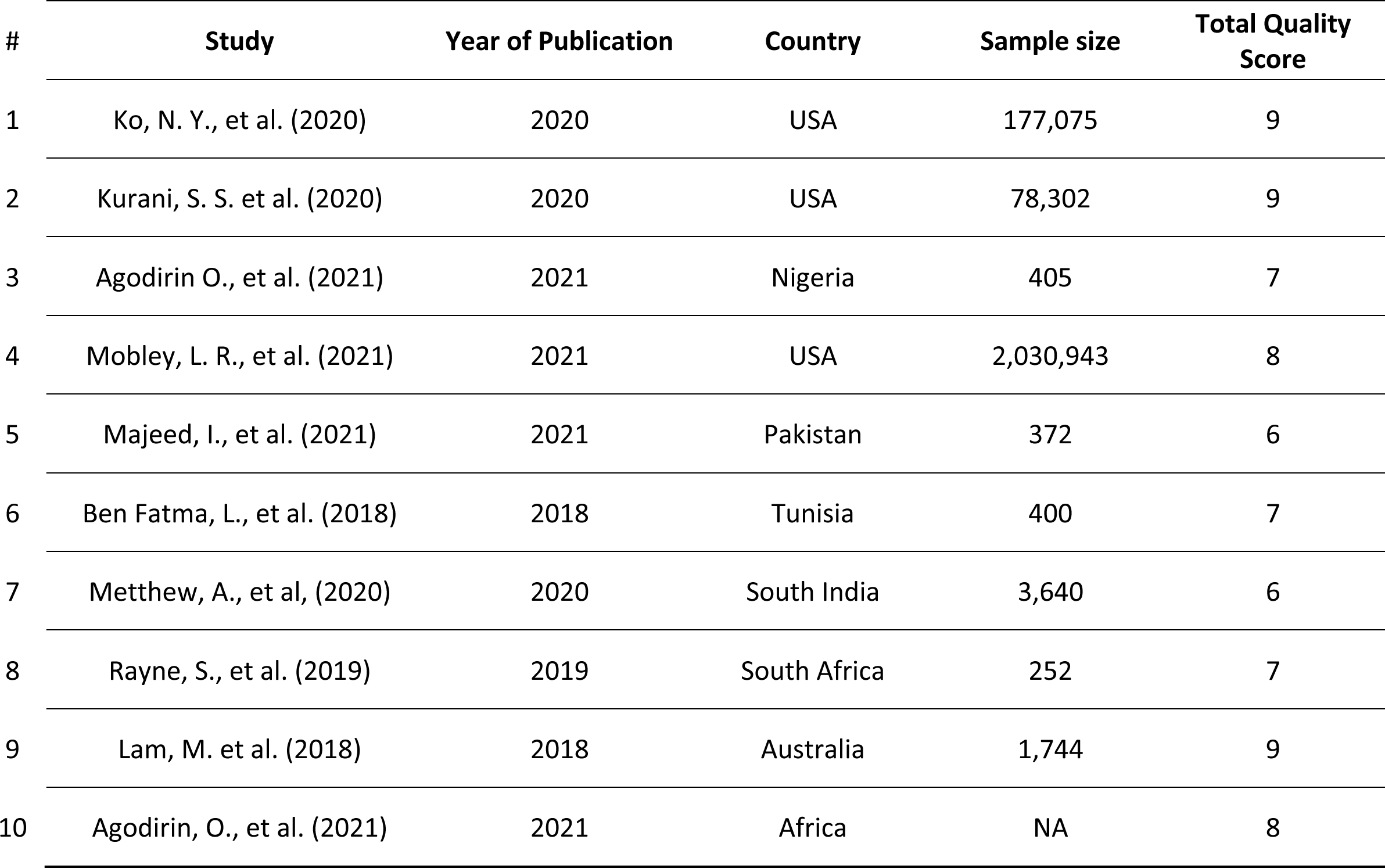
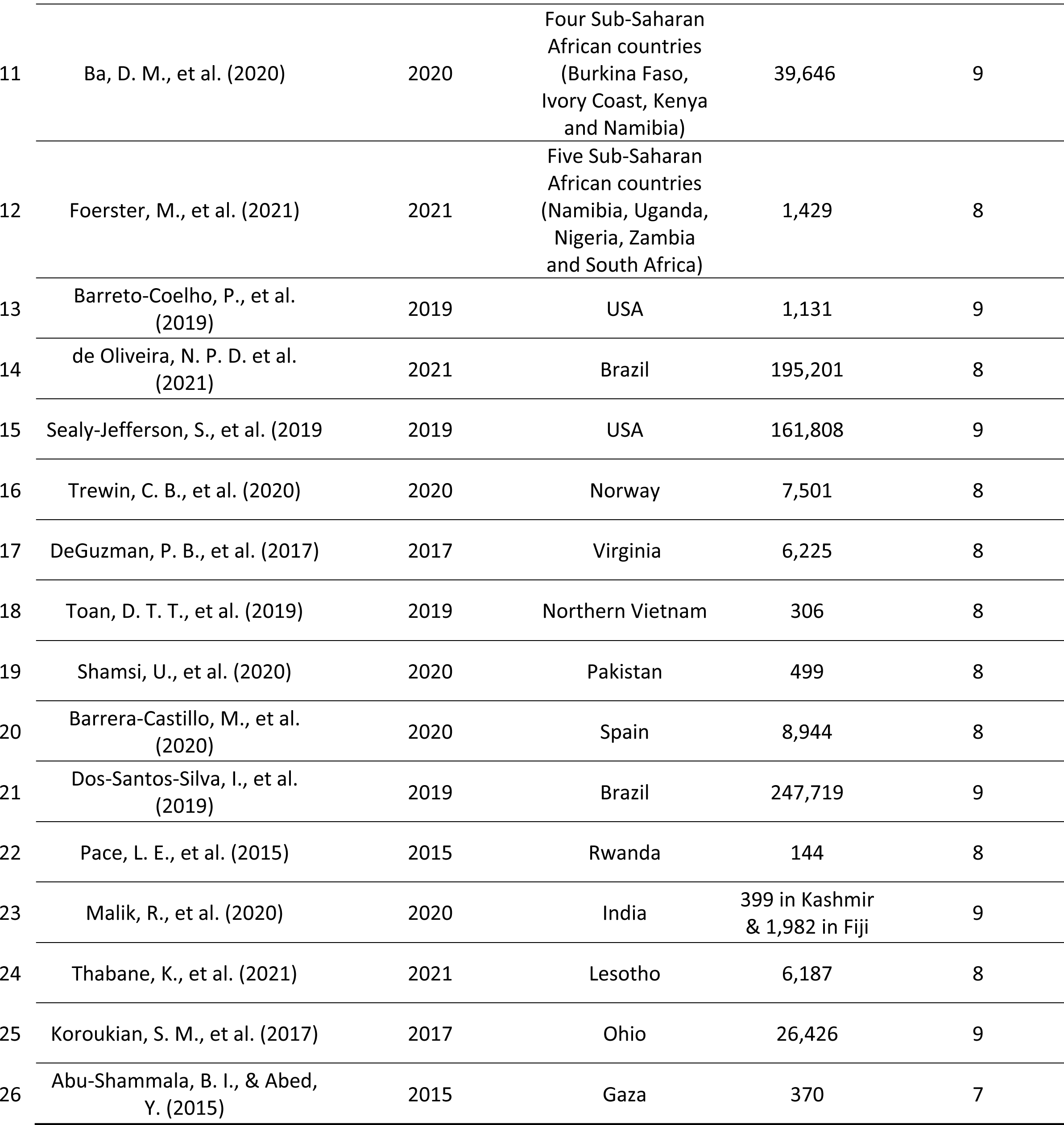
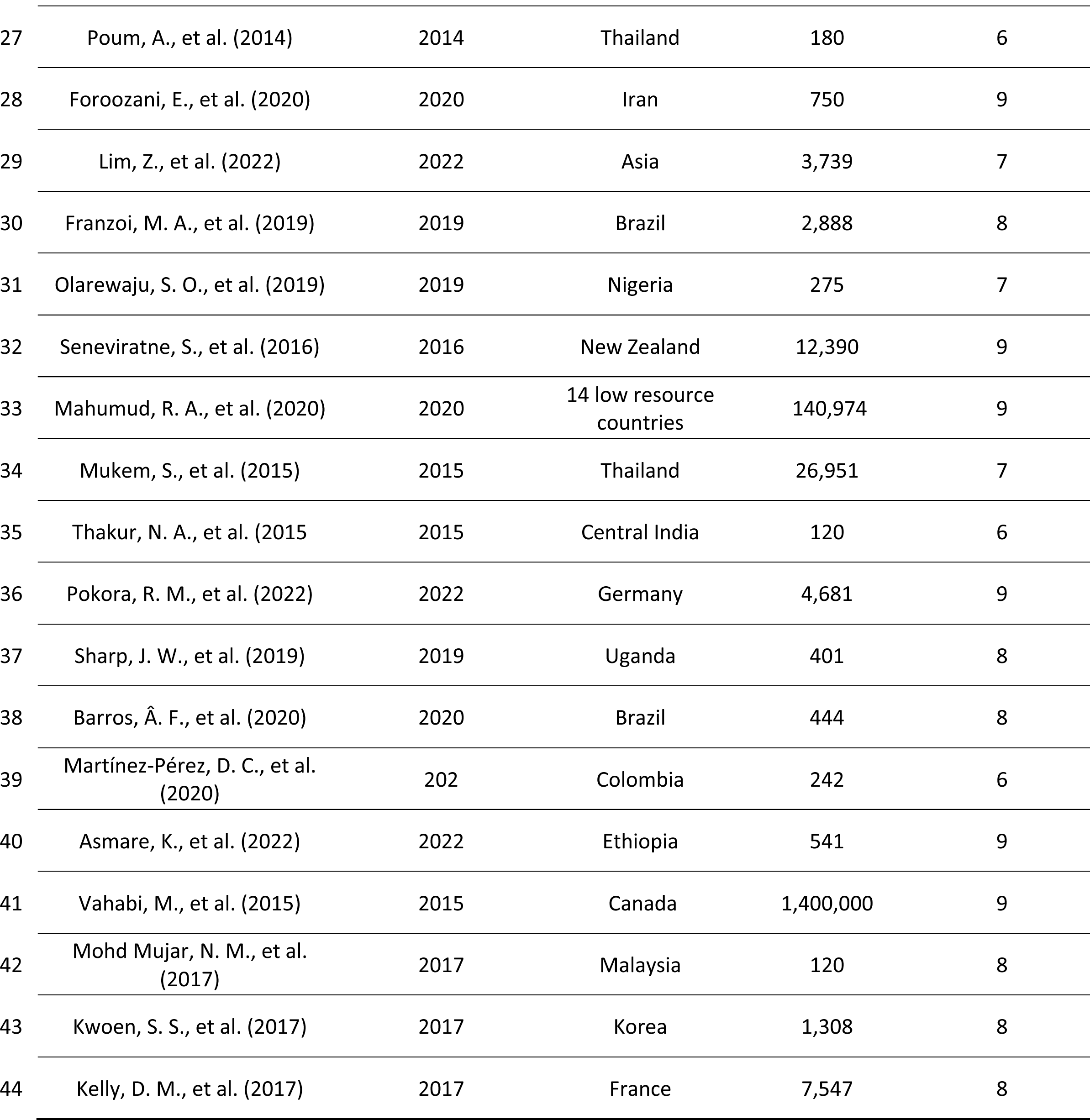
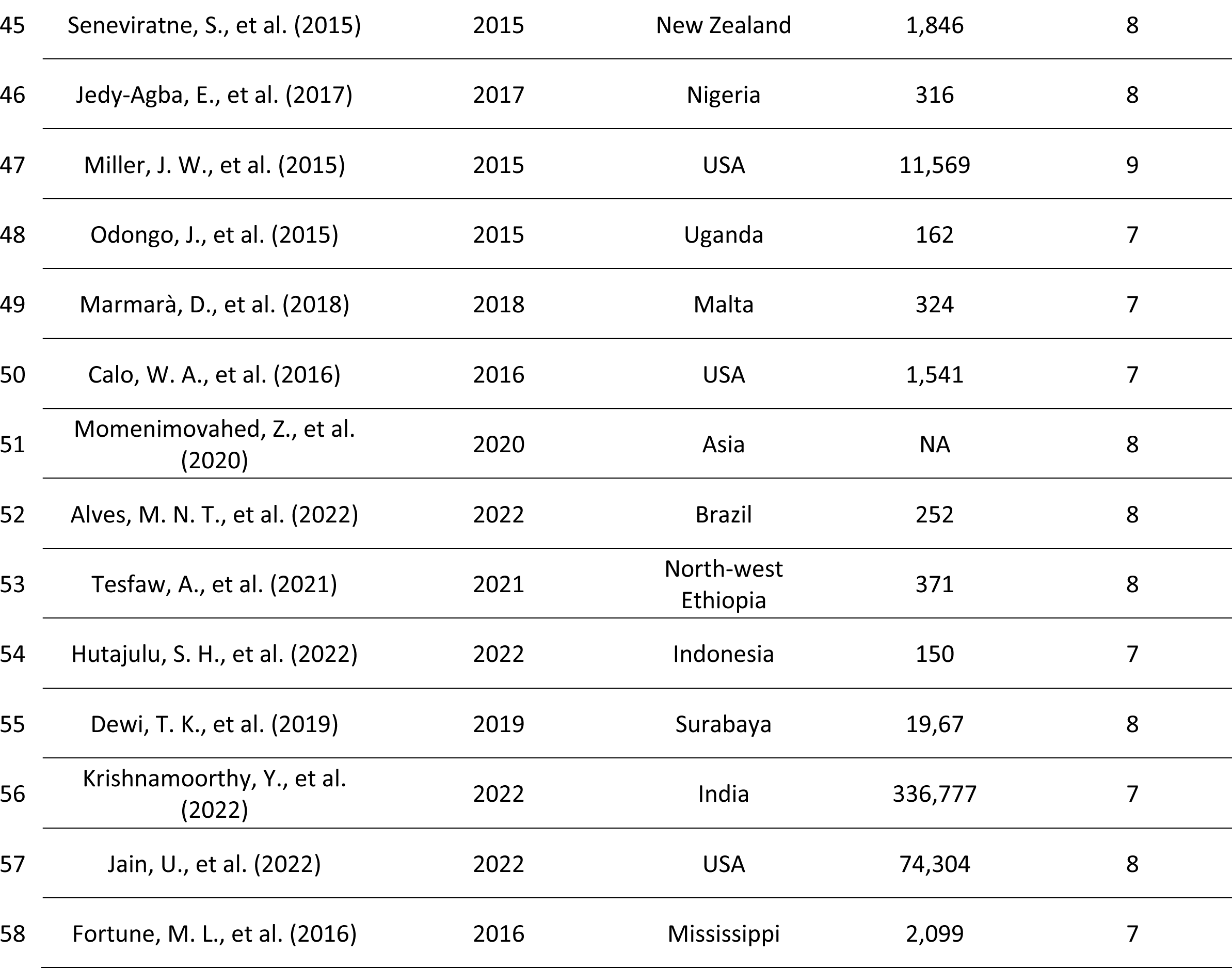
Quality score for the fifty-eight included studies.

